# Handyfuge-LAMP: low-cost and electricity-free centrifugation for isothermal SARS-CoV-2 detection in saliva

**DOI:** 10.1101/2020.06.30.20143255

**Authors:** Ethan Li, Adam Larson, Anesta Kothari, Manu Prakash

**Affiliations:** Department of Bioengineering, Stanford University

**Author notes:** Contributed equally (Ethan Li, Adam Larson).

## Abstract

Point of care diagnostics for COVID-19 detection are vital to assess infection quickly and at the source so appropriate measures can be taken. The loop-mediated isothermal amplification (LAMP) assay has proven to be a reliable and simple protocol that can detect small amounts of viral RNA in patient samples (<10 genomes per μL) (Nagamine, Hase, and Notomi 2002). Recently, Rabe and Cepko at Harvard published a sensitive and simple protocol for COVID-19 RNA detection in saliva using an optimized LAMP assay (Rabe and Cepko, 2020).

This LAMP protocol has the benefits of being simple, requiring no specialized equipment; rapid, requiring less than an hour from sample collection to readout; and cheap, costing around $1 per reaction using commercial reagents. The pH based colorimetric readout also leaves little ambiguity and is intuitive. However, a shortfall in many nucleic acid-based methods for detection in saliva samples has been the variability in output due to the presence of inhibitory substances in saliva. Centrifugation to separate the reaction inhibitors from inactivated sample was shown to be an effective way to ensure reliable LAMP amplification. However, a centrifuge capable of safely achieving the necessary speeds of 2000 RPM for several minutes often costs hundreds of dollars and requires a power supply.

We present here an open hardware solution- *Handyfuge* - that can be assembled with readily available components for the cost of <5 dollars a unit and could be used together with the LAMP assay for point of care detection of COVID-19 RNA from saliva. The device is then validated using the LAMP protocol from Rabe and Cepko. With the use of insulated coolers for reagent supply chain and delivery, the assay presented can be completed without the need for electricity or any laboratory scale infrastructure.

## Methods

We present a design of a new hand powered, low cost centrifuge specifically designed for molecular assay. We evaluated a published LAMP assay for SARS-CoV-2 detection using commercial reagents with this hand powered centrifuge for sample preparation. The assay uses low cost solutions to inactivate, concentrate, and colorimetrically detect small amounts of virus RNA present in human saliva. Our objective was to adapt this assay for use with no external power supply.

To overcome the limitation of requiring a lab centrifuge to remove the inhibitory flocculant we have applied a low cost diagnostic developed in the lab termed the **‘Handyfuge’**. The ‘Handyfuge’ uses a mechanical strategy similar to the ‘Dyno-torch’ flashlights to generate centrifugal force using kinetic input from the user. The user repeatedly squeezes the handle to spin a small freewheel connected to a centrifuge spindle. After a designated amount of time, enough centrifugal force (>500RCF) (Fig. 2) is applied to the eppendorf tube of the inactivated sample to provide inhibitor-free supernatant. This supernatant can then be reliably used for LAMP detection of SARS-CoV-2 or other viruses using the Rabe and Cepko assay.

**Figure 1.**
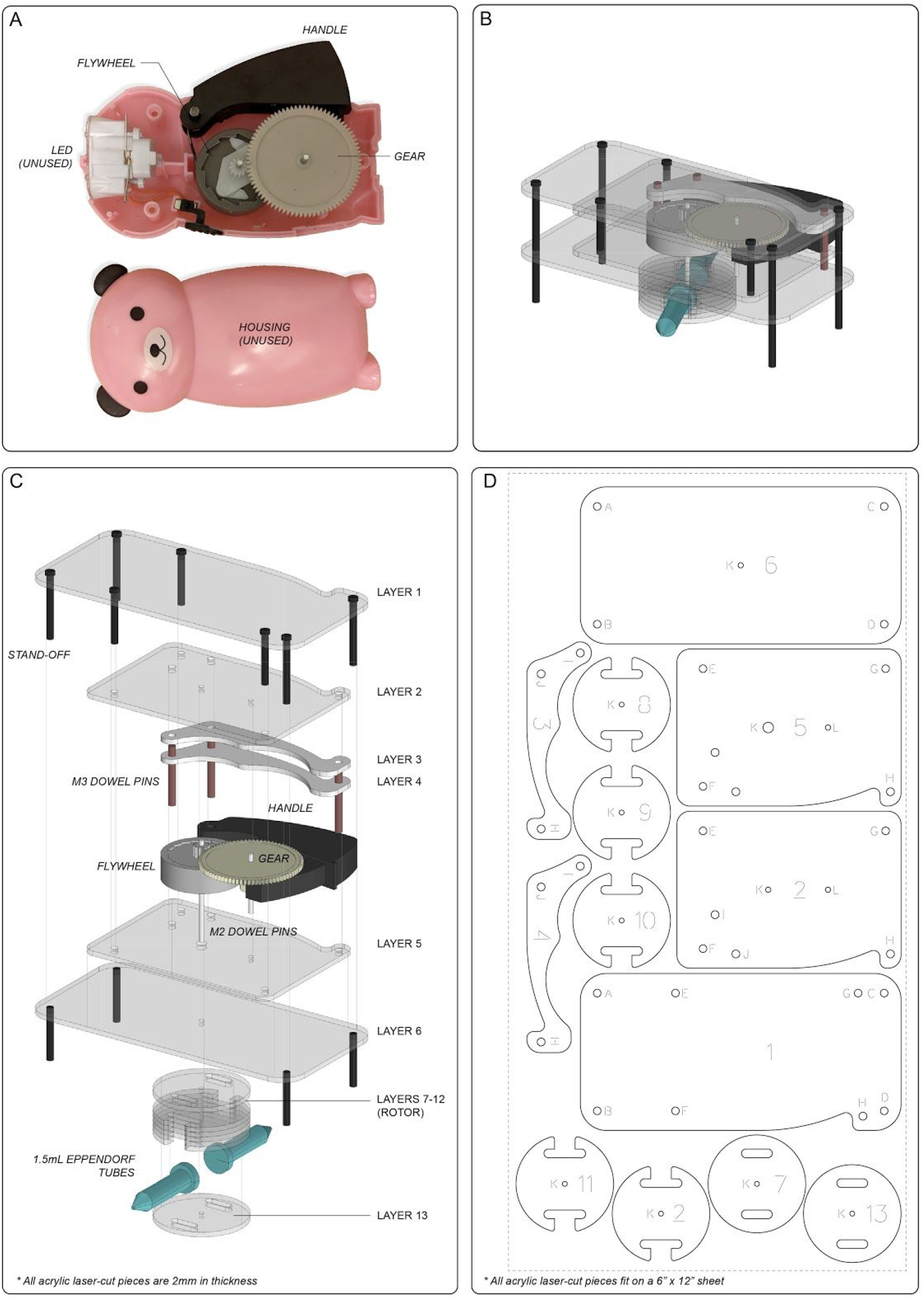
Components of Handyfuge. (A) Dismantled hand-crank flashlight similar to a Dyno-torch flash light providing a one-way ratchet mechanism for converting hand-crank input into uni-directional rotation of the associated gears. (B) Three-dimensional Schematic of a Handyfuge. (C) Assembly and blow-out of of Handyfuge components including borrowed fly-wheel from flash-light and associated acrylic cut pieces. (D) Two-dimensional laser-cut pattern for fabricating a handyfuge.

**Figure 2.**
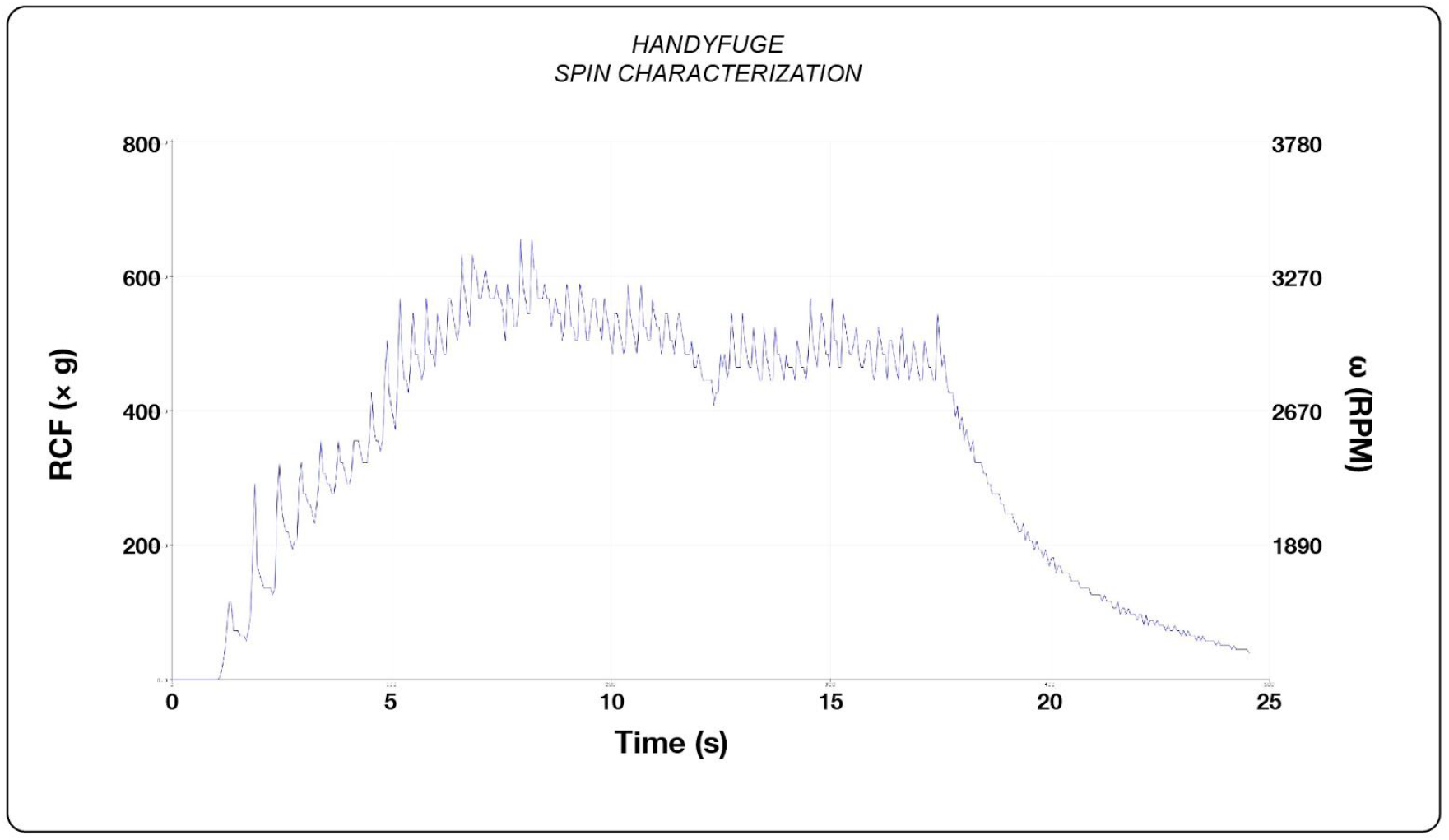
Hall-effect sensor based rotational speed characterization of Handyfuge, demonstrating unidirectional rotation with peak rpm of 3270.

**Modified LAMP protocol** (Adapted from Rabe and Cepko, 2020)

### Reagent Preparation

#### Lysis/Inactivation buffer

To make 1 ml of 100X inactivation reagent, first 71 mg of TCEP-HCl was dissolved in water to create .5 ml of a 0.5 M solution. Then .1 ml of 0.5 M EDTA, pH = 8 was added. Finally, 10 N NaOH and tap water was added to bring the final volume to 1 ml and the NaOH concentration to 1.15N.

#### Chaotropic salt binding solution

To make the NaI binding solution, 45 g of NaI was dissolved in water to a final volume of ∼45 ml. To this, .5 ml of 1 N HCl and 1 ml of TritonX-100 was added and mixed before bringing the volume to 50 ml with tap water.

#### Silica binding solution

To prepare the silica binding solution, 5 grams of 325 mesh silicon dioxide (sigma 342890) was placed in a tube of 50 ml water. After allowing the silica to settle for 1 hour, the supernatant was poured off and another volume of 50 ml water was added. This should clear the solution of silica particles smaller than 1 micron. This can then be mixed with the NaI solution before the assay to make a ‘binding mix’. 5-10 μL per sample is suitable for accurate detection.

Commercial desiccant packets were also used with success, though the variable size range and possibility of contaminants make them less than ideal.

#### LAMP Master Mix

The WarmStart® Colorimetric LAMP 2X Master Mix from New England Biolabs was used as the reaction mix.

#### Primers

Ordered from IDT, sequences from Rabe 2020.

As1_F3

CGGTGGACAAATTGTCAC

As1_B3

CTTCTCTGGATTTAACACACTT

As1_LF

TTACAAGCTTAAAGAATGTCTGAACACT

As1_LB

TTGAATTTAGGTGAAACATTTGTCACG

As1_FIP

TCAGCACACAAAGCCAAAAATTTATCTGTGCAAAGGAAATTAAGGAG

As1_BIP

TATTGGTGGAGCTAAACTTAAAGCCCTGTACAATCCCTTTGAGTG

As1e_FIP T

CAGCACACAAAGCCAAAAATTTATTTTTCTGTGCAAAGGAAATTAAGGAG

As1e_BIP

TATTGGTGGAGCTAAACTTAAAGCCTTTTCTGTACAATCCCTTTGAGTG

#### Mock viral template

Synthetic COVID-19 control RNA (10219) was ordered from Twist and resuspended in nuclease-free water.

## Results

Our initial SARS-CoV-2 assays used saliva spiked with synthetic CoV-2 RNA to validate the efficiency of the Handyfuge in providing viable samples. Preliminary results indicate the LAMP assay after Handyfuge centrifugation is sensitive enough to detect SARS-CoV-2 at less than 100 viral genomes/μL, as reported in the initial publication. The use of the Handyfuge and colorimetric readout allow the assay to be used with nothing more than a heat source capable of heating water or sand to 95°C. A covered ceramic mug and a thermometer is suitable to provide a vessel for inactivation at 95°C, as well as amplification at 65°C. An insulated thermos provides the best results.

### Sample inactivation

The protocol is nearly identical to that published by Rabe and Cepko. Around 500μl of saliva is collected in a microcentrifuge tube. 1/100th sample volume of the 100X TCEP/EDTA solution (inactivation reagent) is then added to the sample. The tube is mixed by inverting then placed in a covered mug of boiling water and incubated for 5 minutes. This step will denature membranes and proteins, while chelating RNase cofactors. The sample is left at neutral pH ∼6.5.

### Centrifugation

At this point the sample has been inactivated through chemical denaturation and heat. However, the saliva still contains inhibitors which will prohibit effective LAMP amplification. To separate the inhibitors, the microcentrifuge tubes containing the inactivated sample are placed in the Handyfuge and spun at max speed for 1 minute by pumping the handle. This is sufficient to separate a precipitate which collects at the bottom of the tube. The cleared supernatant containing inactivated virus is poured into a new tube. This frees the sample from reaction inhibitors present in the saliva flocculant.

### Viral RNA capture

To create a chemical environment amenable to silica/RNA capture, 250 μL of NaI binding reagent is added along with 5 μl of silica slurry to the cleared saliva. The tube is inverted and mixed, then incubated at room temperature for 10 minutes to allow time for the silica particles to interact with the nucleic acid present in the saliva. After this, samples are placed in the Handyfuge and spun again at max speed for 30 seconds, after which the supernatant is poured off. Silica particles are trapped at the bottom of the centrifuge tube.

### Ethanol wash

700 μL of 80% ethanol is added to the sample tube containing the silica and is mixed by inversion. The sample is spun again at max speed 30 seconds, and ethanol is poured off. The residual ethanol then needs to be removed by evaporation. This can be achieved by quickly spinning them to collect the ethanol at the bottom, then placing the tube open into the thermos of hot water to speed up evaporation. Once silica resembles a dry film at the bottom of the tube, the LAMP reaction can proceed.

### LAMP reaction

A 1X mastermix containing the LAMP primers, polymerase, reverse transcriptase, buffers, and indicator dye is added to the tube. The reaction is started by placing the tube at 65°C. After 30 minutes the reaction can be read. A successful polymerization reaction will greatly lower the pH of the solution and turn the indicator dye from purple to yellow.

## Conclusions

The Handyfuge combined with the assay from Cepko and Rabe works reliably detecting synthetic COVID RNA down to 10-100 copies per μl in saliva. To determine the effectiveness as a point of care tool however, it will need to be validated with actual patient samples. We are currently preparing to test this protocol and *Handyfuge* in field settings.

## Materials and Methods

**Figure 3.**
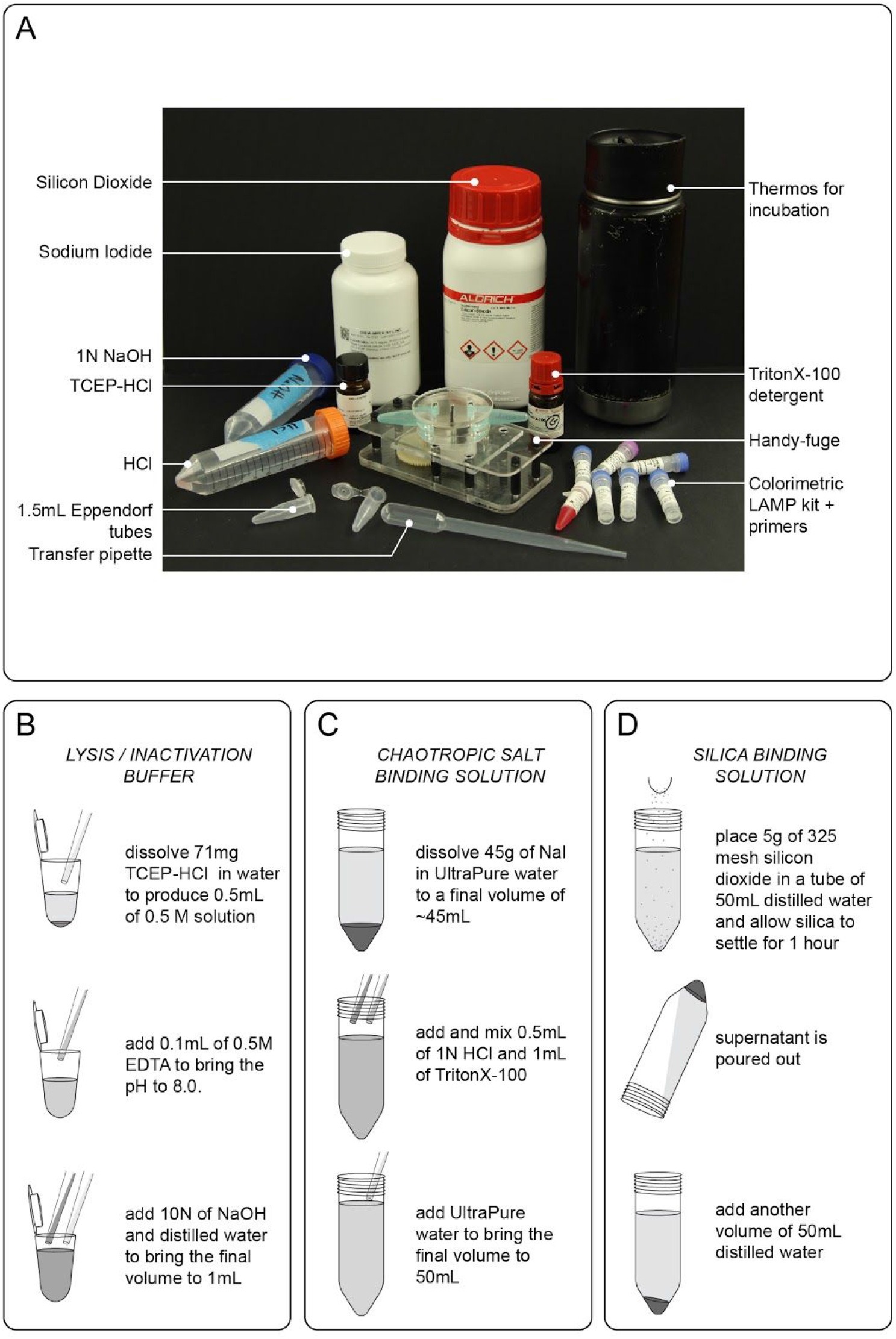
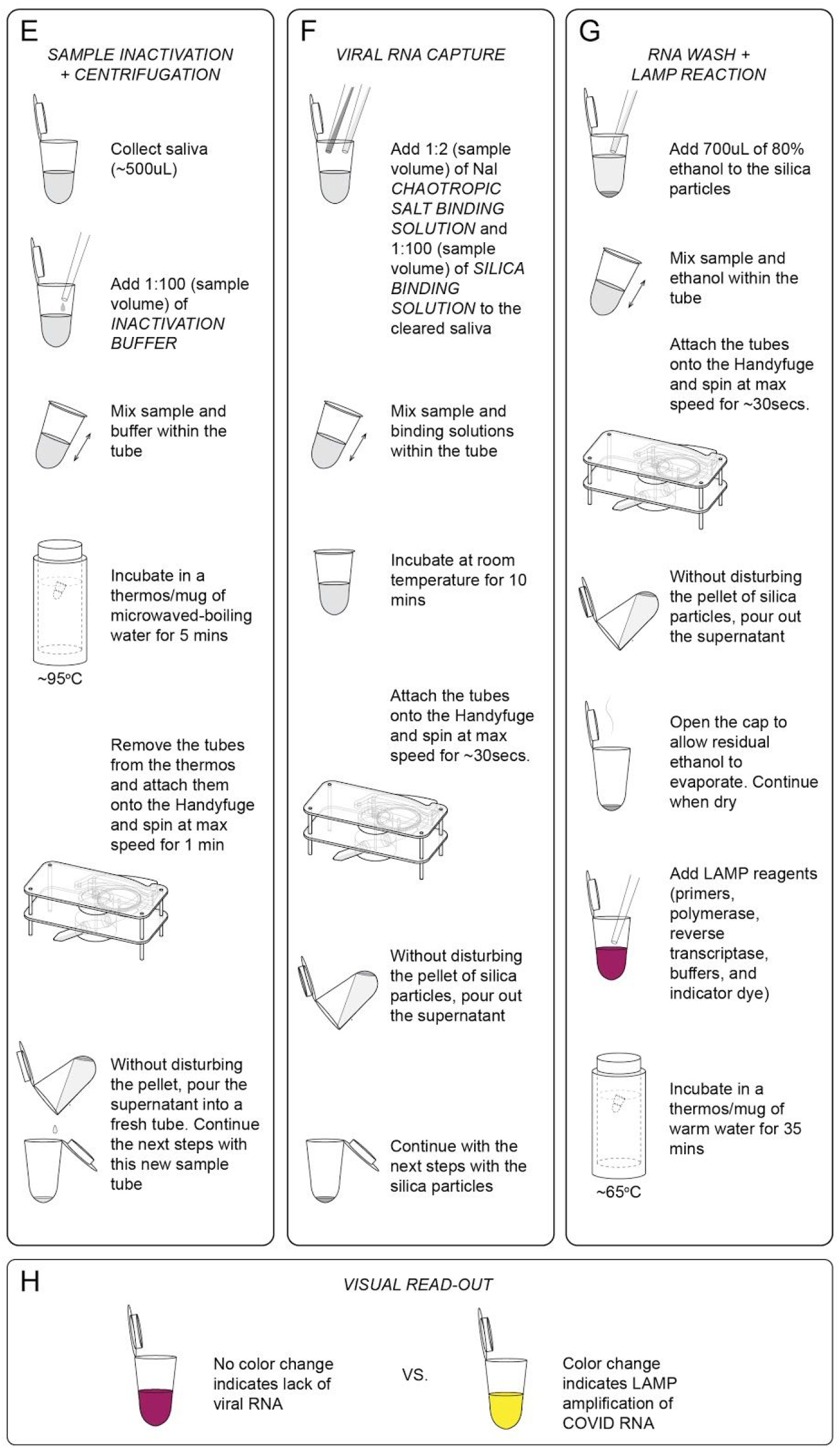
Methods and list of all reagents and steps for Handyfuge-LAMP assay. (A) A picture of all the reagents and tools needed to conduct an electricity-free diagnostics test s. (B)-(D) Preparation of buffers and binding solutions. (E) Sample inactivation (F) Viral RNA capture (G) RNA wash and LAMP reaction. (H) Visual read-out key.

**Figure 4.**
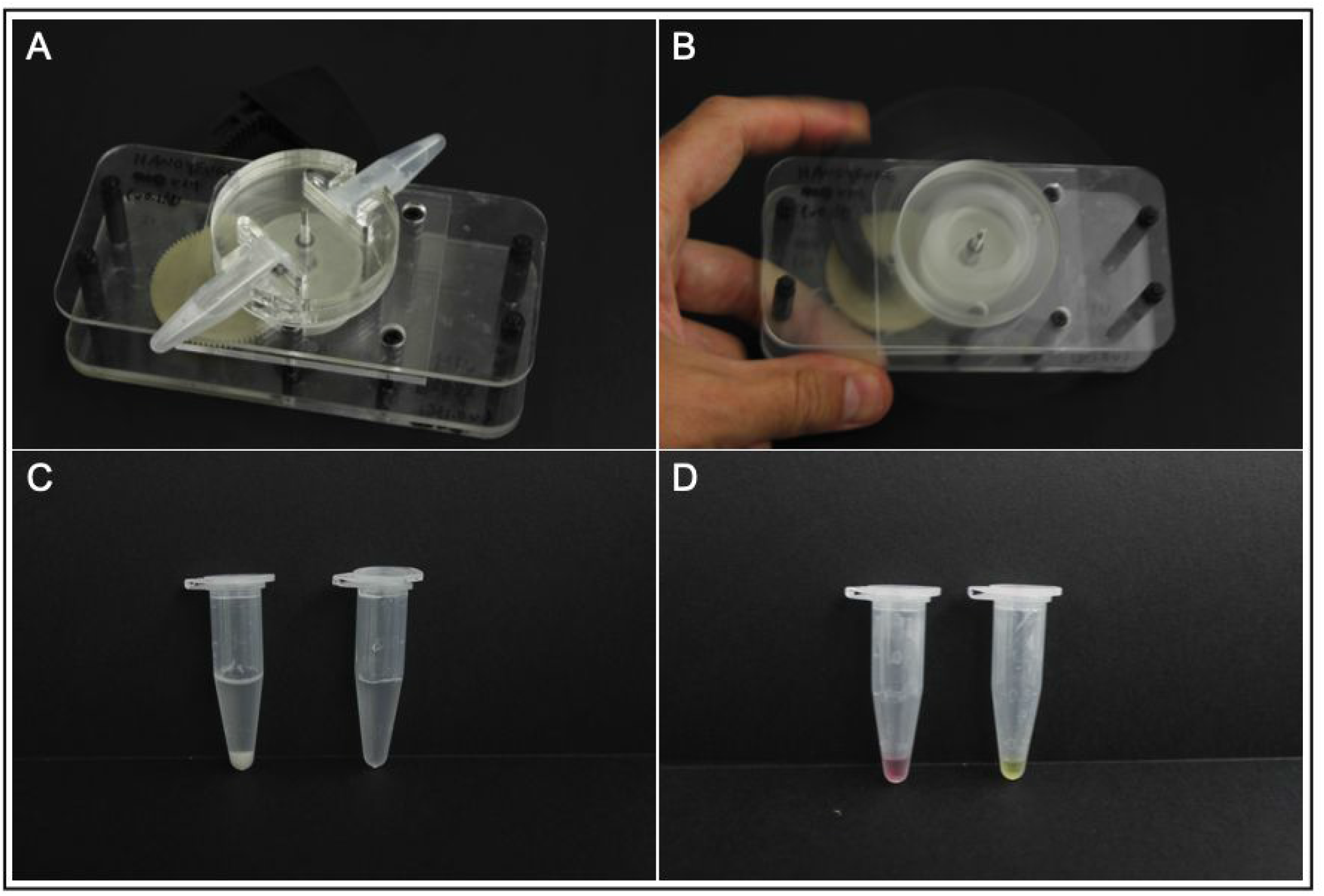
Handyfuge preparation of inhibitor free saliva for LAMP amplification. A) The Handyfuge accepts 2 x 1.5 ml microcentrifuge tubes and a cover for safe operation. B) Hand powered centrifugation can reach sustained speeds of 500RCF. C) Centrifugation at full speed for 1 minute effectively pellets inhibitory flocculant from saliva containing viral RNA. The pelleted flocculant is visible in the tube on the left, the cleared supernatant is on the right. D) After centrifugation, the LAMP protocol can effectively detect synthetic viral RNA down to 10 copies per μl in saliva. The right tube shows the successful amplification of 50,000 COVID control RNA genomes spiked in 500μl of saliva. The tube on the left was spiked with plasmid DNA as a negative control.

**Figure 5.**
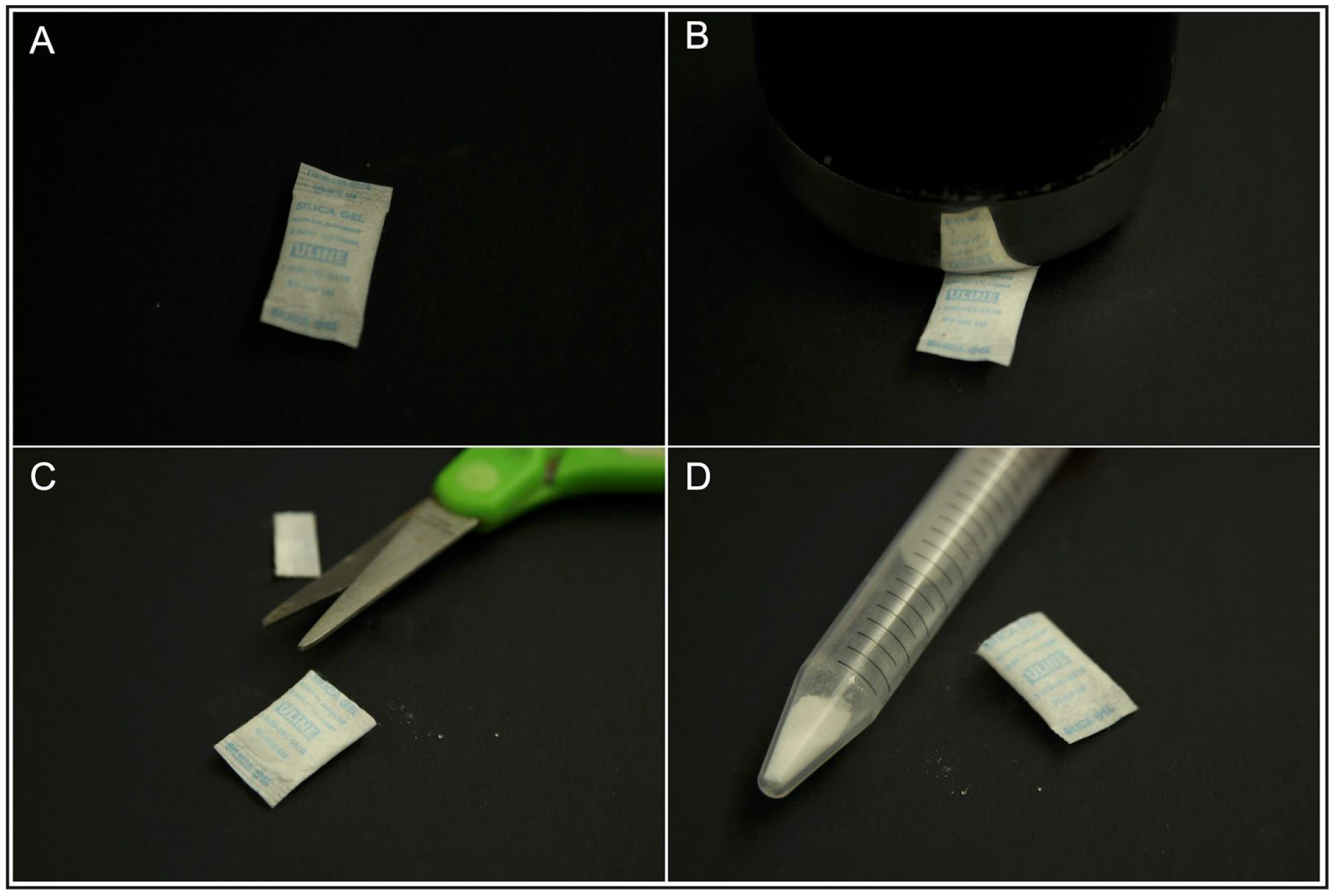
Standard desiccant packs without indicator dye also can be used for RNA capture. A) Standard silica gel desiccant pack. B) The amorphous silica should be pulverised in the package to create a fine powder. Depending on the type of pack the size of the silica ranges from μm to mm. C) After pulverisation the powder can be opened and transferred to a tube. D) The silica will have a much more heterogeneous size range than the normalized lab grade powder, the larger sediments will sink to the bottom of the tube quickly when agitated and the silica remaining in solution can be used for RNA capture.

**Figure 6.**
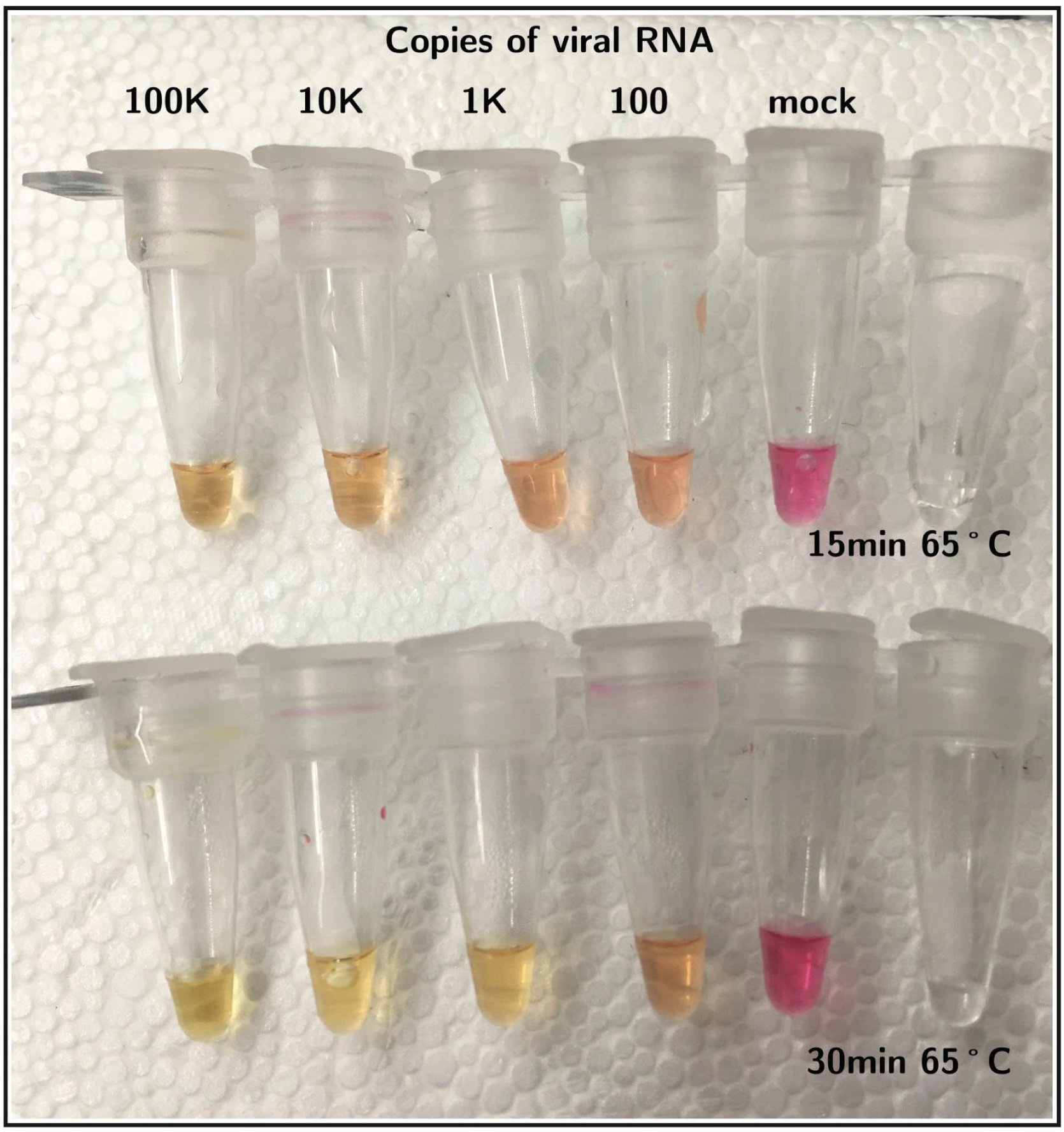
The Handyfuge combined with the Rabe and Cepko protocol detects synthetic COVID RNA at less than 100 copies per μL from saliva. Allowing the reaction to proceed over 30 minutes gives the most unambiguous results.

## Data Availability

All data related to the manuscript is available by request to the authors at.

http://web.stanford.edu/group/prakash-lab/cgi-bin/labsite/covid19/

